# Thoracic Aorta Measurement Extraction from Computed Tomography Radiology Reports Using Instruction Tuned Large Language Models

**DOI:** 10.1101/2024.12.23.24319567

**Authors:** Ely Erez, Sedem Dankwa, McKenzie Tuttle, Afsheen Nasir, Prashanth Vallabhajosyula, Eric B. Schneider, Roland Assi, Chin Siang Ong

**Affiliations:** Division of Cardiac Surgery, Yale School of Medicine, New Haven, CT, USA; Department of Surgery, Yale School of Medicine, New Haven, CT, USA; Harvard T.H. Chan School of Public Health, Boston, MA, USA

## Abstract

Chest computed tomography (CT) is essential for diagnosing and monitoring thoracic aortic dilations and aneurysms, conditions that place patients at risk of complications such as aortic dissection and rupture. However, aortic measurements in chest CT radiology reports are often embedded in free-text formats, limiting their accessibility for clinical care, quality improvement and research purposes. In this study, we developed a multi-method pipeline to extract structured aortic measurements from radiology reports, and compared the performance of fine-tuned BERT-based models with instruction-tuned Llama large language models (LLMs). Applying the best-performing method to a real-world large chest CT radiology report database, we generated a comprehensive aortic measurement dataset that facilitates big data aortic disease research.

## Introduction

Chest computed tomography (CT) is essential for diagnosing and monitoring thoracic aortic dilations and aneurysms. These conditions, often asymptomatic, significantly increase the risk of life-threatening complications such as aortic dissection and rupture^1^. Frequently, thoracic aortic dilations are detected incidentally on chest CTs performed for unrelated clinical indications. Once diagnosed, chest CTs serve as the preferred modality for tracking aortic diameter changes over time and guiding surgical decision-making^1^. However, key diagnostic measures, including aortic diameters, are predominantly documented in free-text CT radiology reports, rendering them inaccessible in a structured format within most electronic health record systems.

Extracting these measures in a structured form could make a substantial clinical impact. These structured data can be used to flag reports with incidental findings within the EHR software, drawing the attention of physicians and facilitating timely referrals to surgeons. This can prevent missed findings, improve the quality of care by monitoring referral rates, and adapt to evolving evidence-based medicine (EBM) guidelines that adjust surgical referral thresholds. Additionally, healthcare institutions can monitor surgical referral rates and ensure adherence to the most current EBM, enhancing patient outcomes and healthcare quality. From the research perspective, extracting these measures retrospectively in a large cohort of patients will enable researchers to study rates of incidental aortic dilation detection and analyze how aortic diameters evolve over time in affected patients. This structured data can also provide valuable epidemiological insights into the prevalence and risk factors associated with thoracic aortic dilations.

The task of extracting aortic diameters falls within the broader domain of information extraction, which is commonly addressed using natural language processing (NLP) techniques. Specifically, it requires word- or token-level classification, akin to named entity recognition (NER), where specific entities are identified and categorized within text. NER techniques have evolved rapidly in recent years, transitioning from hand-crafted rule-based approaches to feature-driven statistical models and, ultimately, to end-to-end deep learning models^2^. As a key application of NER, Biomedicine has been a central area of research^3^, with extensive work exploring its use in medical imaging^4^. Early studies relied on hand-crafted rules to identify entity mentions, often involving complex logic and requiring substantial domain expertise^5,6^. Later work employed more advanced classical machine learning techniques such as Hidden Markov Models^7^, Support Vector Machines^8^, and Conditional Random Fields^9^. While these methods improved performance, they still depended on manual feature extraction, a process heavily reliant on domain knowledge, as a prerequisite to classification.

The advent of deep neural networks marked a paradigm shift in NER. Models such as Recurrent Neural Networks (RNNs) and Bidirectional Long Short-Term Memory (BiLSTM) networks achieved state-of-the-art performance while eliminating the need for manual feature engineering, enabling end-to-end learning directly from raw data. These models, in turn, were rapidly surpassed by pretrained language models like the Bidirectional Encoder Representations from Transformers (BERT) model, which leverage encoder-based transformer architectures to set new benchmarks in NER tasks. BERT and its domain specific variants, such as PubMedBERT^10^ and BioBERT^11^, have become the gold standard in NER. By fine-tuning on prelabeled datasets, these models achieve high performance in NER tasks with minimal reliance on additional domain knowledge. These models have been widely adopted by the medical community for information extraction^3^, including significant effort to improve information extraction from radiology reports^4^. For instance, Khurshid et al.^12^ developed Bio+Discharge Summary BERT, a fine-tuned BERT model for NER, to extract vital signs such as height, weight, and blood pressure from unstructured electronic health record (EHR) notes, reducing vital sign data missingness by 31%. Similarly, Singh et al.^13^ addressed a challenge closely related to ours, using a fine-tuned BERT model to extract 21 quantitative measures from cardiac magnetic resonance imaging. Their approach achieved a macro-average F1 score of 0.957, highlighting the strong performance of these models with minimal labeling effort.

In recent years generative LLMs such as OpenAI’s GPT-3.5 and GPT-4^14^, have transformed the field of natural language processing, breaking performance records across numerous benchmarking tasks. These models utilize a Transformer architecture scaled to hundreds of billions of parameters, enabling unprecedented contextual understanding and fluency in text generation. The introduction of smaller, open-source LLMs such as Meta’s Llama^15^, expanded research opportunities by making these technologies accessible to the research community enabling the development of tools for fine-tuning and inference on more modest hardware. The ability of pretrained LLMs to perform zero-shot^16^ and few-shot^17^ learning has allowed them to excel in many NLP tasks without extensive labeled data. Recognizing this potential, researchers have sought to apply similar approaches to biomedical NER^18,19^, hoping to reduce reliance on labeled training data while maintaining competitive performance.

In practice, pretrained LLMs have struggled to match the performance of fine-tuned BERT-based models on NER tasks^20,21^. Instruction tuning, in contrast, has shown far greater potential^22,23^. Instruction tuning, or instruction fine-tuning, describes the process by which large language models undergo supervised fine-tuning to better follow instructions and improve performance on a given task^24^. An example of this technique is a study by Keloth et al.^25^, who developed BioNER-Llama 2 by instruction-tuning Llama 2-7B on three publicly available biomedical NER datasets focusing on diseases, chemicals and genes. BioNER-Llama 2 achieved performance comparable to fine-tuned PubMedBERT, with F1 scores ranging from 0.949 to 0.956 on the test sets. Similarly, Bian et al.^26^ created the VANER model by instruction-tuning Llama 2-7B on eight biomedical NER datasets, reporting F1 scores between 0.77 and 0.94, consistent with fine-tuned BERT-based models. Notably, both VANER and BioNER-Llama 2 demonstrated poor generalizability, with significantly reduced performance on previously unseen datasets. Additionally, these studies primarily focus on benchmark tasks, which, while useful for assessing baseline capabilities, may not reflect real-world complexities. he real-world performance of LLMs in NER applications, particularly in the biomedical domain, remains largely underexplored.

The primary objective of this study was to develop an automated machine learning pipeline for extracting aortic measurements from chest CT radiology reports. To achieve this, we compared the performance of fine-tuned BERT-based models with generative LLMs. A secondary objective was to construct a comprehensive aortic measurement database by applying the pipeline to a large cohort of chest CT radiology reports from our institution. The generated dataset will enable future investigations into patterns of aortic dilation detection and the progression of aortic disease in a hospital-based population.

## Methodology

This study was determined to be exempt from review by Yale University’s Institutional Review Board (IRB) under protocol number 2000037866 on May 3, 2024.

### Dataset

We conducted an institutional search for chest CTs with corresponding radiology reports for patients aged 18 and older, performed between January 2013 and December 2023. The search encompassed 43 distinct CT protocols and yielded 363,423 radiology reports. Reports from CT protocols with fewer than 2,000 instances and reports without narrative content were excluded, resulting in a final dataset of 356,690 radiology reports across 16 CT protocols.

A subset of 1,506 radiology reports was selected for manual annotation using stratified random sampling to ensure balanced representation across protocols. This subset was divided into 1,002 reports for training (sampled at a 1:356 ratio) and 504 reports for validation and testing (sampled at a 1:712 ratio), ensuring that reports in the training set were distinct from the reports used for validation and testing to prevent information leakage. The narratives were annotated using Label Studio^27^ by two medical students and a postdoctoral researcher with an MD. To ensure consistency, all annotations were reviewed by the postdoctoral researcher. Thoracic aortic diameters were labeled at eight anatomical sites: the annulus, sinus of Valsalva, sinotubular junction, mid ascending, ascending proximal to the brachiocephalic, top of the arch, proximal descending, and mid descending.

Due to the limited number of aortic diameter annotations identified in the 504 reports intended for validation and testing (n=289 annotations), the reports designated for both validation and testing were instead allocated exclusively for validation. To create the testing set, an additional 504 reports were sampled and annotated following the same protocol. Reports selected for training and validation were excluded from the pool when selecting the test set to avoid data leakage. This process resulted in 2,010 labeled reports, divided into training, validation, and testing sets with a 50:25:25 split. Figure 1 provides a flowchart illustrating the dataset selection process, while Table 1 presents the radiology reports and corresponding patient characteristics for each set.

**Figure 1:**
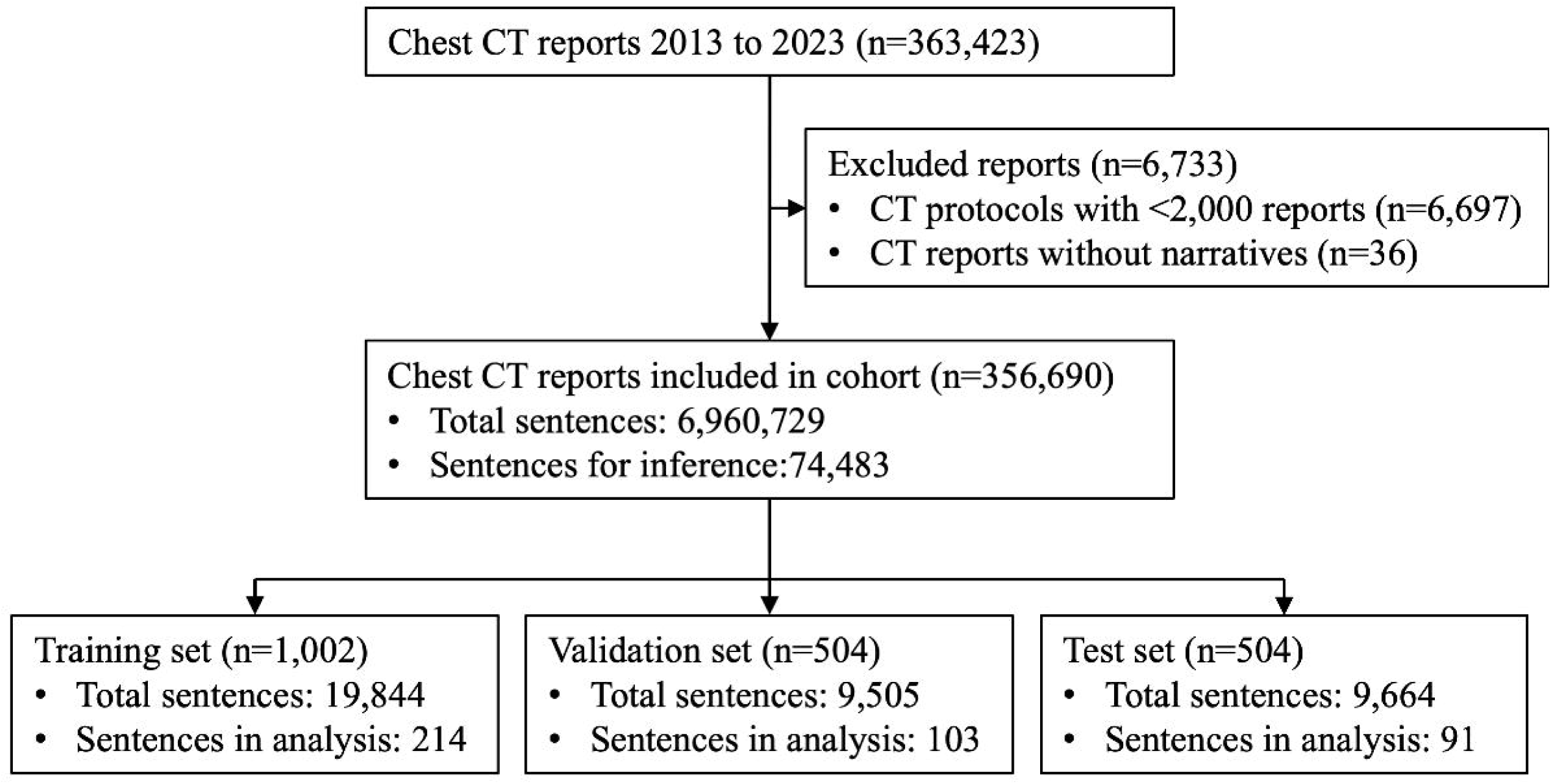
Flowchart illustrating the selection and annotation process of radiology reports, divided into training, validation, and testing sets.

**Table 1.** Train, validation and test dataset radiology report characteristics.

|  | **Train** | **Validation** | **Test** |
| --- | --- | --- | --- |
| Radiology reports (N) | 1002 | 504 | 504 |
| CT type (n [%]) |  |  |  |
| CT chest without IV contrast | 288 [28.7] | 144 [28.6] | 144 [28.6] |
| CT chest with IV contrast | 196 [19.6] | 98 [19.4] | 98 [19.4] |
| CTA chest (PE) with IV contrast | 172 [17.2] | 86 [17.1] | 86 [17.1] |
| CT chest, abdomen, pelvis with IV contrast | 157 [15.7] | 79 [15.7] | 79 [15.7] |
| CT ED chest, abdomen, pelvis with IV contrast | 27 [2.7] | 14 [2.8] | 14 [2.8] |
| CTA chest, abdomen, pelvis with and/or without IV contrast | 24 [2.4] | 12 [2.4] | 12 [2.4] |
| CT chest, abdomen, pelvis without IV contrast | 22 [2.2] | 11 [2.2] | 11 [2.2] |
| CTA chest with and/or without IV contrast | 18 [1.8] | 9 [1.8] | 9 [1.8] |
| CT chest without IV contrast, high resolution | 17 [1.7] | 9 [1.8] | 9 [1.8] |
| CTA chest, abdomen with and/or without IV contrast | 16 [1.6] | 8 [1.6] | 8 [1.6] |
| CTA coronary | 16 [1.6] | 8 [1.6] | 8 [1.6] |
| CT initial lung cancer screening | 12 [1.2] | 6 [1.2] | 6 [1.2] |
| CT cardiac scoring without IV contrast | 12 [1.2] | 6 [1.2] | 6 [1.2] |
| CT subsequent lung cancer screening | 11 [1.1] | 6 [1.2] | 6 [1.2] |
| CT thoracic spine without IV contrast | 7 [0.7] | 4 [0.8] | 4 [0.8] |
| CTA chest vascular with and/or without IV contrast/gated | 7 [0.7] | 4 [0.8] | 4 [0.8] |
| Age (median [IQR]) | 66 [55-75] | 65 [54-75] | 66 [54-76] |
| Females (n [%]) | 490 [48.9] | 272 [54.0] | 259 [51.4] |
| Race |  |  |  |
| White | 778 [77.6] | 384 [76.2] | 382 [75.8] |
| Black or African American | 117 [11.7] | 67 [13.3] | 78 [15.5] |
| Asian | 11 [1.1] | 4 [0.8] | 8 [1.6] |
| American Indian or Native American | 1 [0.1] | 1 [0.2] | 2 [0.4] |
| Native Hawaiian or Other Pacific Islander | 2 [0.2] | 3 [0.6] | 1 [0.2] |
| Other | 67 [6.7] | 31 [6.2] | 28 [5.6] |
| Missing | 20 [2.0] | 12 [2.4] | 5 [1.0] |

### Data preprocessing

To account for BERT’s token limit and ensure a fair comparison with Llama models, we opted to perform fine-tuning and inference on individual sentences rather than complete narratives. Because most sentences in the report narratives did not contain aortic measurements, splitting the reports into sentences allowed us to exclude irrelevant content, thereby improving label balance and significantly reducing fine-tuning and inference times. Sentence splitting was performed using the Python package NLTK. The first sentence of each report, as well as any sentence lacking a non-time or date numeric value, was excluded from analysis. Additionally, we retained only sentences containing at least one aorta-related keyword.

For fine-tuning BERT-based models, each aortic measurement site was assigned a numeric label between 1 and 8. Tokens within the span of a measurement were assigned the corresponding numeric label, while all other tokens were labeled as 0. This numerical vector served as the label for each sentence. For Llama models we used XML tags to delineate measurements in the target output, such as <SOV> and</SOV> for the sinus of Valsalva. If no measurements were present, the output remained identical to the input sentence. This annotation approach, similar to that employed by Keltoh et al.^25^, facilitates straightforward postprocessing. Figure 2 shows a sample input and output for Llama models.

**Figure 2:**
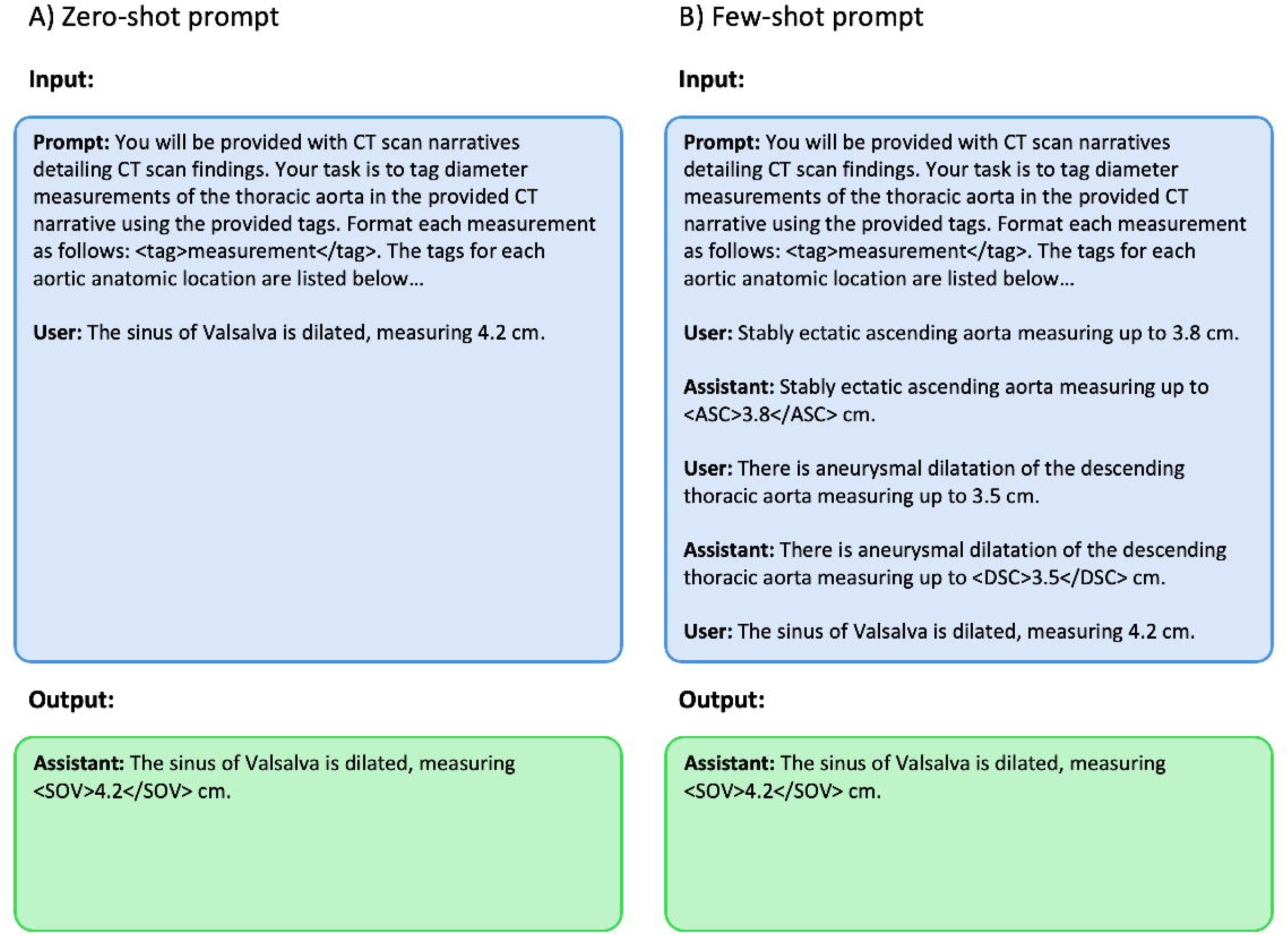
Sample input and output. (A) Zero-shot prompt and (B) Few-shot prompt for Llama models, illustrating the use of XML tags to delineate aortic measurement sites.

### Baseline Model

We used Meta’s Llama 3.1 instruction-tuned model with 8 billion (8B) parameters as a baseline model^28^. This 8B model was chosen for its strong benchmark performance relative to its size, which represents the upper limit of our virtual machine’s capacity for local fine-tuning. The instruction-tuned version was chosen over the base (pre-trained) version because it underwent several rounds of alignment, including supervised fine-tuning, rejection sampling, and direct preference optimization, which improved its instruction-following capability, quality, and safety^29^. The baseline model’s performance was further optimized through prompt engineering, following the methodology described by Hu et al.^19^. The prompt included a task description, labeling instructions based on the annotation guidelines, and additional instructions informed by error analysis conducted on the training set. We evaluated the baseline model’s zero-shot performance as well as few-shot performance using three pre-selected annotated samples from the training set. The selected prompt, as well as several annotated radiology report samples, are publicly available on GitHub at https://github.com/yalesurgeryresearch/RadTextExtractor.

### BERT-based Models

We fine-tuned six BERT-based models by combining three weight initialization strategies with two tokenization schemes. The weight initialization strategies included: (1) the original BERT model^30^, (2) BERT-NER^31^, a variant fine-tuned on the English CoNLL-2003 Named Entity Recognition dataset^32^, and (3) PubMedBERT^10^, pre-trained on PubMed abstracts and full-text articles from PubMed Central. The two tokenization schemes tested were: (1) the standard BERT tokenization and (2) a modified scheme in which numeric values were replaced with the unique [NUM] pseudo-token^33^.

Fine-tuning was performed using the AutoModelForTokenClassification class from the HuggingFace transformers library^34^, which adds a token classification head to the BERT architecture for mapping hidden states to output labels. The fine-tuning process adhered to the baseline scheme proposed by Mosbach et al.^35^. Each model was fine-tuned on the selected sentences from the training set over 20 epochs, using categorical cross-entropy loss. Optimization was conducted with the AdamW^36^ optimizer, employing a linear learning rate scheduler and a 10% warm-up phase. Hyperparameter tuning was conducted using grid search, testing learning rates of 1e-5, 2e-5, 5e-5 and 1e-4, along with batch sizes of 8, 16, and 32. A random seed was fixed during training to ensure reproducibility. Loss on the validation set was calculated after each epoch, and the epoch with the lowest validation loss was selected for each fine-tuning iteration.

### Llama Models

We compared the performance of three versions of Meta’s Llama models using instruction-tuning: the Llama 2 chat-tuned model with 7 billion parameters, the Llama 3 instruction-tuned model with 8 billion parameters, and the Llama 3.1 instruction-tuned model with 8 billion parameters. To accommodate the instruction-tuning process on our virtual machine’s limited GPU RAM, we employed 4-bit Quantized Low-Rank Adaptation (QloRA)^37^. QLoRA builds on Low-Rank Adaptation (LoRA)^38^, a technique that significantly reduces the number of trainable parameters, by further quantizing model weights to 4-bit precision. This approach enables fine-tuning of large language models on resource-constrained hardware while maintaining high performance. Additionally, we utilized Unsloth^39^, an open-source library that accelerates fine-tuning through a custom backpropagation engine.

Instruction-tuning for each model used the same prompt as the zero-shot baseline Llama 3.1 model. Instruction-tuning was conducted on sentences from the training set over 5 epochs, using cross-entropy loss and an AdamW^36^ optimizer with a linear rate scheduler without a warm-up phase. A batch size of 1 was used for all trials. During fine-tuning, we zeroed-out the loss on the provided prompt, ensuring learning only on the model’s output. Validation loss was calculated after each epoch, and the epoch with the lowest validation loss was selected for each instruction-tuning trial. Hyperparameter tuning was performed using grid search to optimize the learning rate, as well as LoRA’s rank, and alpha parameters. The learning rates tested were 2e-5, 5e-5, and 1e-4. Rank values included 16, 32, and 64, with alpha values set to rank multiplied by 1 or 2 (e.g., for a rank of 16, the alpha values tested were 16 and 32). Sampling was disabled during inference to ensure deterministic and reproducible results. When labeling sentences, the model was provided with the prompt and a sentence as input and generated a labeled output.

### Evaluation metrics

Model performance was evaluated on the validation set using exact match macro-averaged precision, recall, and F1 scores across all aortic measurement sites, as well as site-specific precision, recall, and F1 scores. To simplify terminology, macro-averaged precision, recall, and F1 scores are referred to as “macro precision,” “macro recall,” and “macro F1,” respectively. Performance metrics were calculated across the entire validation set, including sentences not selected for inference during preprocessing. The optimal model from the baseline models, BERT-based models, and Llama models was selected based on the highest macro F1 score on the validation set.

### Ablation study

To evaluate the impact of training set size on model performance, we conducted an ablation study in which the optimal model was fine-tuned using subsets of 10, 25, 50, and 100 randomly selected sentences from the training set. To mitigate the effects of sentence selection, this process was repeated five times, each time using a different random seed to generate distinct sentence subsets. The median macro F1 score across the five trials was then compared to the performance of the model trained on the full training set.

Following the ablation study, we evaluated the optimal model on the test set to assess its generalizability and potential real-world performance. Finally, inference was conducted on the entire radiology report cohort to create an aortic measurement database. This process was limited to sentences selected according to the criteria outlined in the preprocessing phase. Measurement extraction rates, aortic dilation rates, and aortic diameter median and interquartile range (IQR) at each measurement site were analyzed as supplementary indicators of the model’s real-world performance.

### Computational Resources and Framework

All analyses were conducted on a HIPAA-compliant virtual machine hosted by Yale’s Spinup service, utilizing Amazon Elastic Cloud Compute (EC2). The environment comprised an Amazon AWS G4 instance with 4 vCPUs, 16 GB of RAM, and an NVIDIA T4 GPU with 16 GB of GPU memory. GPU acceleration was facilitated using CUDA (v12.4). Fine-tuning and inference were performed using Python (v3.9.13) with the following key libraries: PyTorch (v2.3.0), Hugging Face Transformers (v4.43.3), and Unsloth (v2024.8).

The code for data preprocessing, model fine-tuning, and evaluation is available in a GitHub repository at https://github.com/yalesurgeryresearch/RadTextExtractor. Due to the presence of protected health information in the radiology reports, the datasets generated and analyzed in this study, along with the fine-tuned models, are not publicly available. However, they can be obtained from the corresponding author upon reasonable request, in accordance with institutional policies and any applicable data access agreements.

## Results

Following preprocessing, the training dataset used for fine-tuning consisted of 214 out of 19,844 sentences (1.08%), of which 166 (77.6%) contained at least one annotation. The training set included a total of 589 annotations, with a median of 52 annotations per measurement site (range: 44 to 166). The validation set contained 103 out of 9,505 sentences (1.08%) selected for inference, of which 71 (68.9%) included at least one annotation. All sentences with annotations in the validation set were included in the inference subset, which had a total of 289 annotations, with a median of 25.5 annotations per measurement site (range: 20 to 76). The test set comprised 91 out of 9,664 sentences (0.94%) selected for inference, with 68 (74.7%) containing at least one annotation. Similar to the validation set, all sentences with annotations in the test set were included in the inference subset, which contained 215 annotations, with a median of 18 annotations per measurement site (range: 13 to 69). A summary of the annotation characteristics for the training, validation, and test datasets after preprocessing is provided in Table 2.

**Table 2.**
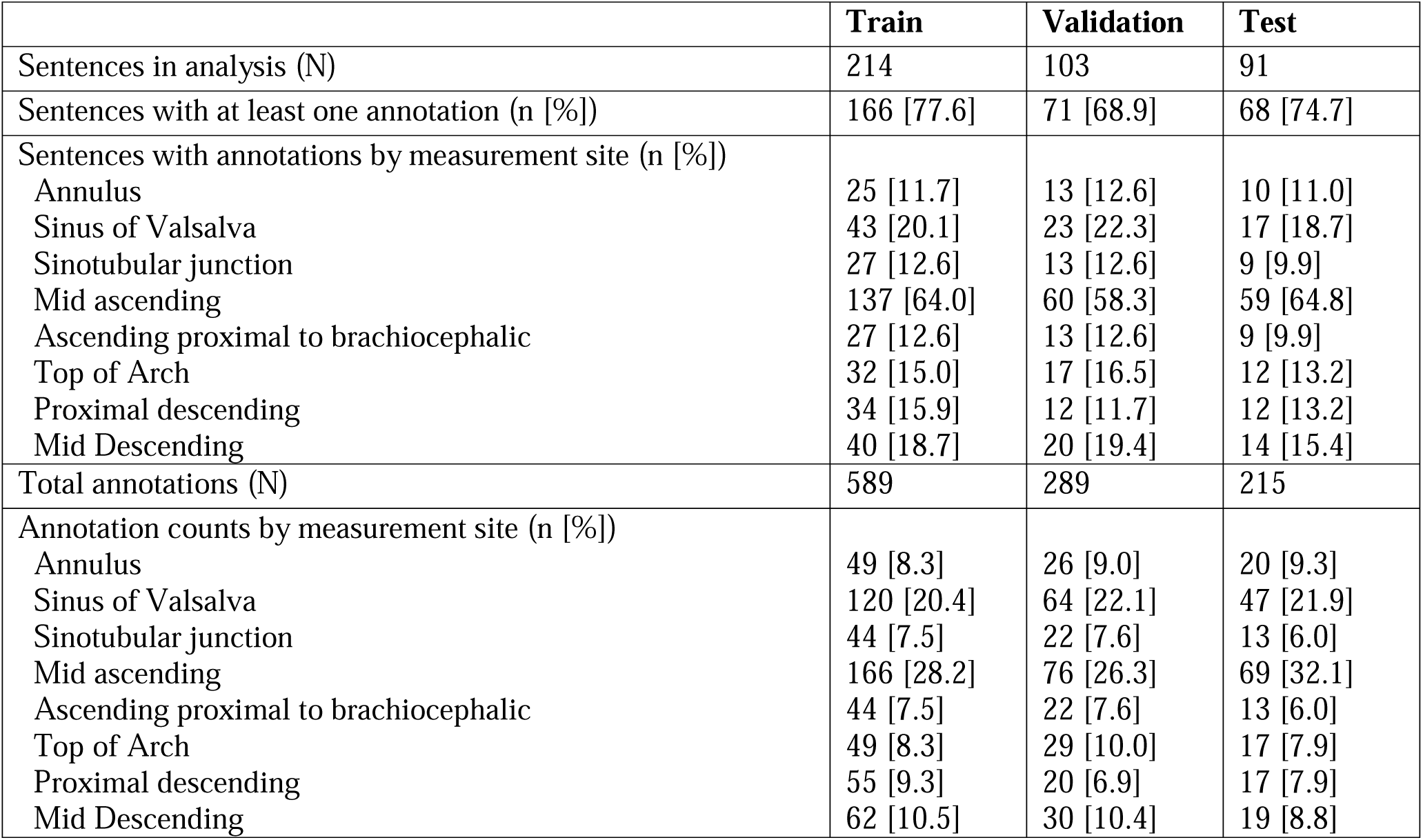
Train, validation and test dataset annotation characteristics following preprocessing.

### Model Performance Comparison

The performance of the baseline Llama 3.1 models, fine-tuned BERT-based models, and instruction-tuned Llama models on the validation set is summarized in Table 3. The few-shot Llama 3.1 was the best performing baseline model, besting the zero-shot model with a macro F1 score of 0.838 compared to 0.663. However, both baseline models were surpassed by all fine-tuned BERT-based models and instruction-tuned Llama models.

**Table 3.**
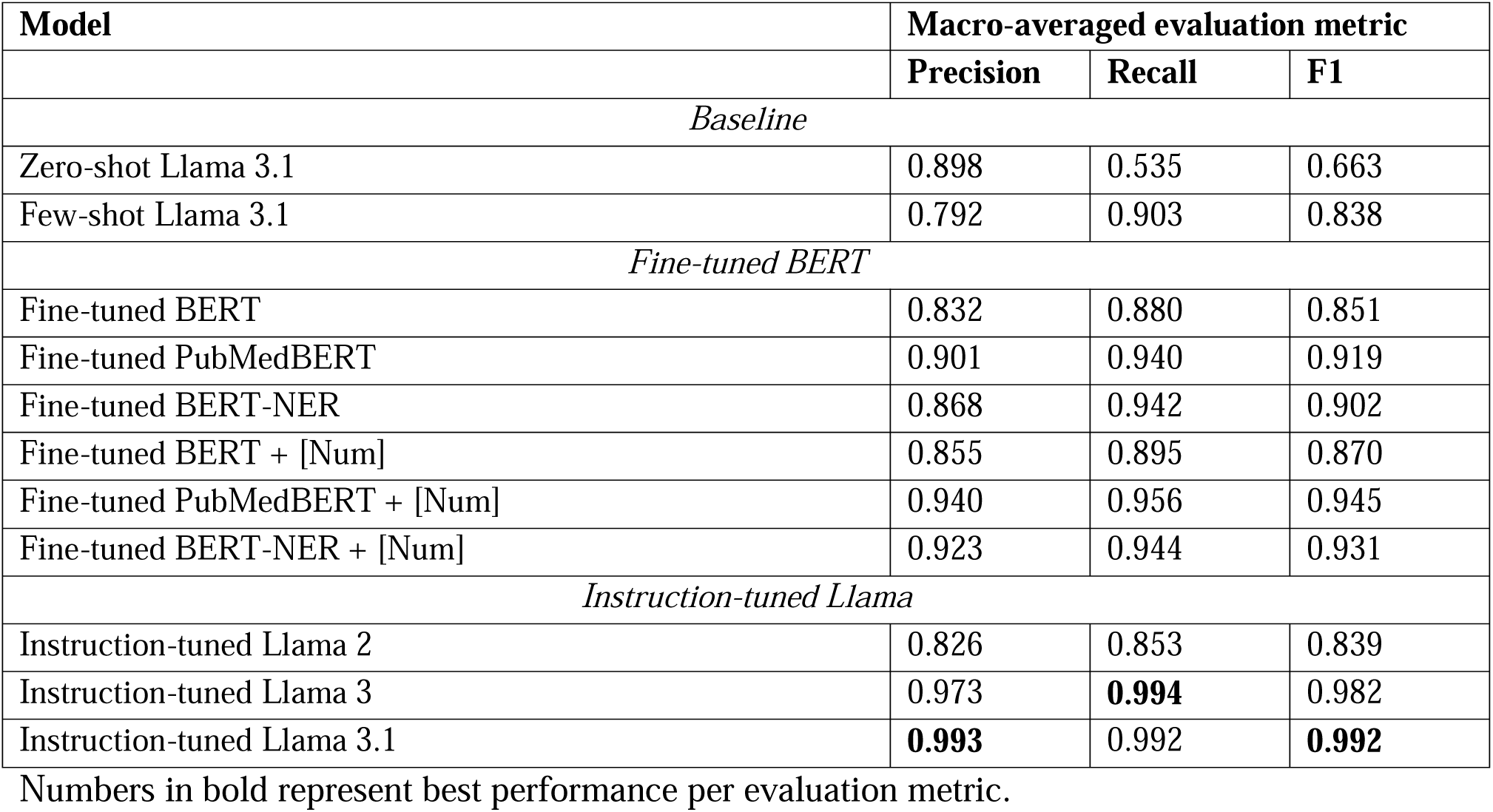
Comparison of model performance on validation set.

| Model | Macro-averaged evaluation metric |  |  |
| --- | --- | --- | --- |
|  | Precision | Recall | F1 |
| *Baseline* |  |  |  |
| Zero-shot Llama 3.1 | 0.898 | 0.535 | 0.663 |
| Few-shot Llama 3.1 | 0.792 | 0.903 | 0.838 |
| *Fine-tuned BERT* |  |  |  |
| Fine-tuned BERT | 0.832 | 0.880 | 0.851 |
| Fine-tuned PubMedBERT | 0.901 | 0.940 | 0.919 |
| Fine-tuned BERT-NER | 0.868 | 0.942 | 0.902 |
| Fine-tuned BERT + [Num] | 0.855 | 0.895 | 0.870 |
| Fine-tuned PubMedBERT + [Num] | 0.940 | 0.956 | 0.945 |
| Fine-tuned BERT-NER + [Num] | 0.923 | 0.944 | 0.931 |
| *Instruction-tuned Llama* |  |  |  |
| Instruction-tuned Llama 2 | 0.826 | 0.853 | 0.839 |
| Instruction-tuned Llama 3 | 0.973 | **0.994** | 0.982 |
| Instruction-tuned Llama 3.1 | **0.993** | 0.992 | **0.992** |

Numbers in bold represent best performance per evaluation metric.

The best performance among the BERT-based models was achieved by the fine-tuned PubMedBERT with [NUM] tokenization, which attained a macro F1 score of 0.945. Numeric tokenization using a [NUM] pseudo-token consistently outperformed the standard tokenization technique across all three models, and the fine-tuned PubMedBERT was the best performing model in both the standard and [NUM] tokenization.

The instruction-tuned Llama 3.1 delivered the highest overall performance, achieving a near-perfect macro F1 score of 0.992, with a macro precision of 0.993 and macro recall of 0.992. The instruction-tuned Llama 3 followed closely with a macro F1 score of 0.982. Notably, the instruction-tuned Llama 2 achieved a macro F1 of 0.839, performance comparable to the few-shot baseline model and significantly lower than the top-performing BERT-based models.

Figure 3 compares the distributions of F1 scores across measurement sites for the few-shot Llama 3.1, PubMedBERT with [NUM] tokenization, and the instruction-tuned Llama 3.1. The few-shot Llama 3.1 and PubMedBERT models showed significant variability in F1 scores across different measurement sites. In contrast, the instruction-tuned Llama 3.1 demonstrated consistent performance, achieving an F1 score of at least 0.971 across all sites.

**Figure 3:**
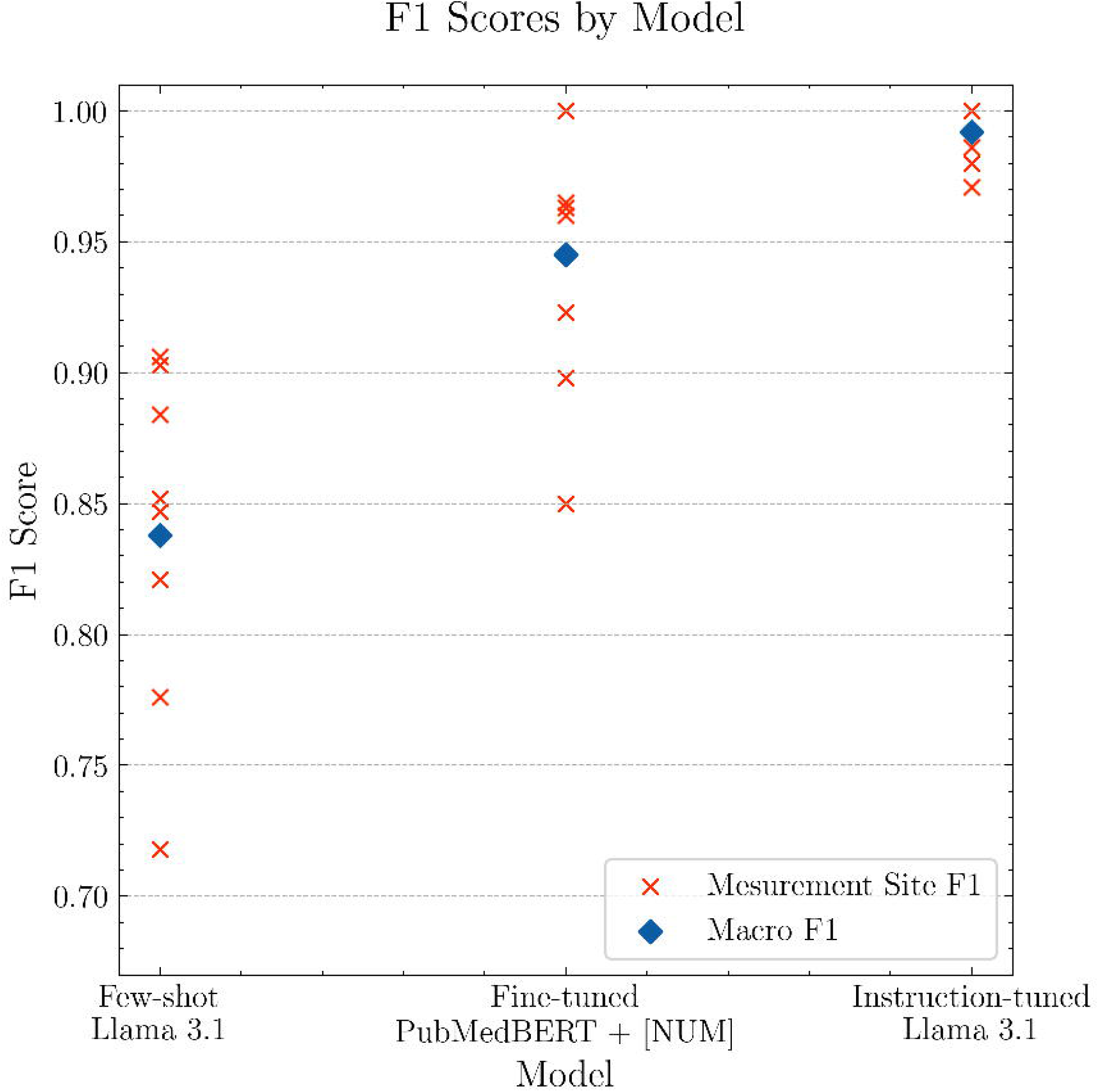
F1 Scores by Model. F1 scores for few-shot Llama 3.1, finetuned PubMedBERT + [NUM], and instruction-tuned Llama 3.1.

### Impact of Training Set Size

Figure 4 shows the results of the ablation study, with macro F1 scores of the instruction-tuned Llama 3.1 model as a function of the number of training sentences. Performance improved rapidly with the initial increase in training sentences but slowed and eventually plateaued as the number grew. Median macro F1 scores were 0.800 with 10 training sentences, 0.880 with 20, 0.903 with 50, and 0.971 with 100, compared to 0.992 when using the full training set of 214 sentences. The results also highlight significant variability in performance due to the random selection of sentence subsets, which decreased as the training set size increased.

**Figure 4:**
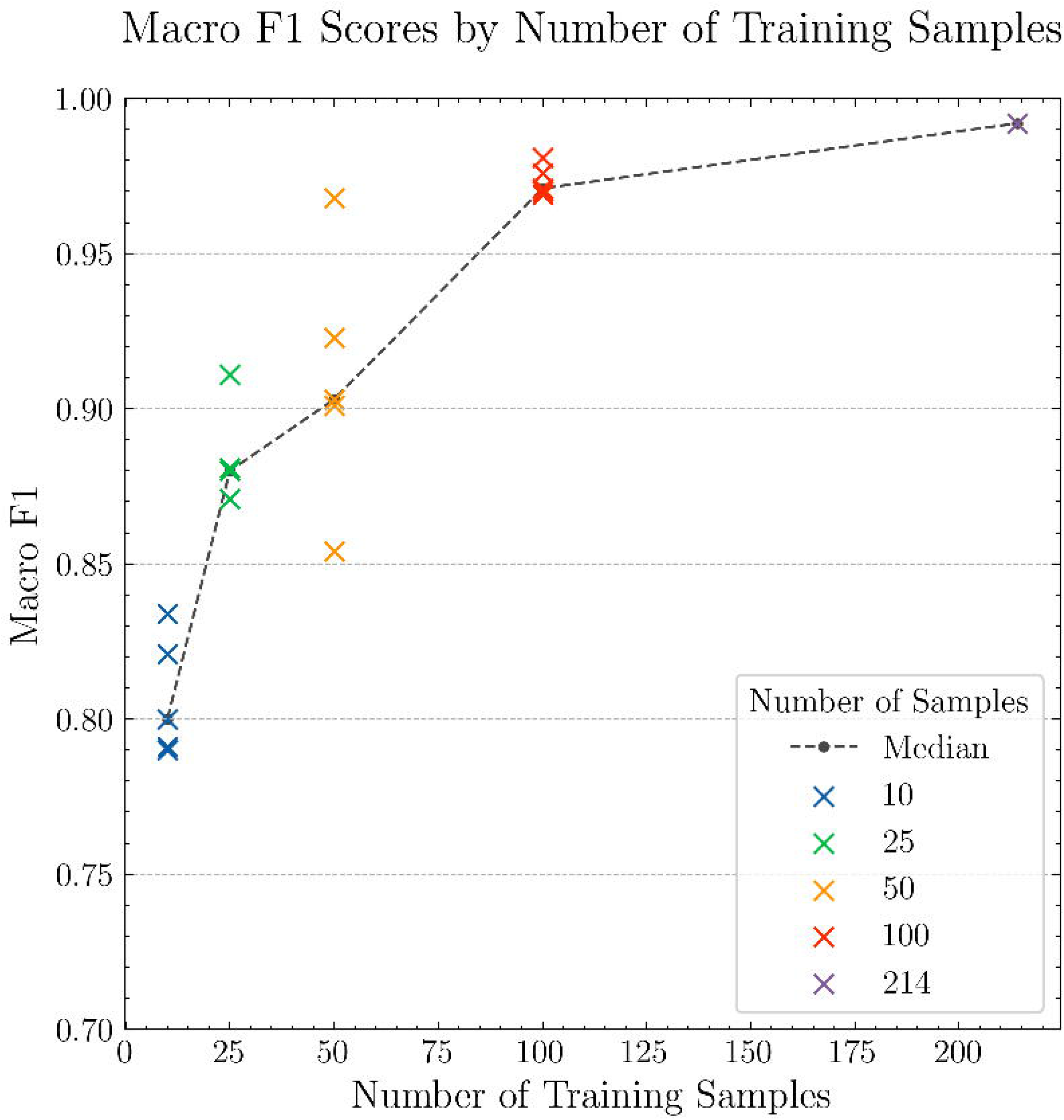
Ablation Study. Macro F1 scores by number of training samples: 10, 25, 50, 100, 214.

### Assessing Model Generalizability

The performance of the instruction-tuned Llama 3.1 model was evaluated on the test set to assess its generalizability to unseen data sampled from the same source distribution. Table 4 compares the model’s performance across aortic measurement sites between the validation and test sets. The macro F1 score on the test set was 0.970, slightly lower than the 0.992 achieved on the validation set. Measurement site F1 scores ranged from 0.923 to 1.000 on the test set, compared to 0.971 to 1.000 on the validation set.

**Table 4.** Fine-tuned Llama 3.1 performance on validation and test sets by aortic measurement site.

| Measurement site | Validation set |  |  | Test set |  |  |
| --- | --- | --- | --- | --- | --- | --- |
|  | Precision | Recall | F1 | Precision | Recall | F1 |
| Annulus | 1.000 | 1.000 | 1.000 | 1.000 | 1.000 | 1.000 |
| Sinus of Valsalva | 1.000 | 1.000 | 1.000 | 1.000 | 1.000 | 1.000 |
| Sinotubular junction | 1.000 | 1.000 | 1.000 | 0.857 | 1.000 | 0.923 |
| Mid ascending | 1.000 | 0.973 | 0.986 | 1.000 | 0.985 | 0.992 |
| Ascending proximal to brachiocephalic | 1.000 | 1.000 | 1.000 | 0.929 | 1.000 | 0.963 |
| Top of arch | 1.000 | 0.962 | 0.980 | 1.000 | 1.000 | 1.000 |
| Proximal descending | 0.944 | 1.000 | 0.971 | 0.938 | 0.938 | 0.938 |
| Mid descending | 1.000 | 1.000 | 1.000 | 0.900 | 1.000 | 0.947 |
| **Macro- averaged** | **0.993** | **0.993** | **0.992** | **0.953** | **0.990** | **0.970** |

Numbers in bold represent best performance per evaluation metric.

### Insights from Full Dataset Extraction

The complete chest CT radiology report dataset consisted of 356,690 reports from 140,645 unique patients. Following preprocessing, 74,483 sentences out of 6,960,729 (approximately 1.07%) were selected for extraction, consistent with the proportions observed in the labeled datasets. After extraction using the instruction-tuned Llama 3.1, 49,387 radiology reports (13.85%) contained at least one aortic measurement, showing higher rates of aortic measurement reporting in males (18.51%) compared to females (9.50%). Table 5 summarizes the extraction results by aortic measurement site. Measurement extraction rates across the aortic sites were similar to those in the labeled datasets (Table 2).

**Table 5.** Complete dataset inference results by aortic measurement site.

| Measurement Site | Reports with measurements (n) | Percent of sentences analyzed with measurements (%) | Max Diameter (mm, median [IQR]) |
| --- | --- | --- | --- |
| Annulus | 7,072 | 9.49% | 28 [25.7-30.3] |
| Sinus of Valsalva | 12,620 | 16.94% | 36 [32-40] |
| Sinotubular junction | 7,621 | 10.23% | 34 [30-37] |
| Mid ascending | 44,472 | 59.71% | 39 [36-42] |
| Ascending proximal to brachiocephalic | 8,056 | 10.82% | 35 [32-39] |
| Top of arch | 9,865 | 13.24% | 31 [28-34] |
| Proximal descending | 8,886 | 11.93% | 30 [27-34] |
| Mid descending | 13,939 | 18.71% | 29 [25-34] |

The largest median diameters were observed at the mid ascending aorta and the sinus of Valsalva, measuring 39 mm (IQR 36–42) and 36 mm (IQR 32–40), respectively. Median diameters decreased distally along the aorta, measuring 31 mm (IQR 28–34) at the aortic arch, 30 mm (IQR 27–34) at the proximal descending aorta, and 29 mm (IQR 25–34) at the mid descending aorta. Ascending aortic dilation of at least 40 mm was reported in 8.69% of patients (12,228/140,645), with 2.27% (3,193/140,645) reported to have a dilation of at least 45 mm, 0.66% (925/140,645) at least 50 mm, and 0.28% (393/140,645) at least 55 mm.

## Discussion

In this study, we describe our experiences developing a machine learning pipeline for extracting aortic measurements from chest CT radiology reports. Among the models evaluated, the instruction-tuned Llama 3.1 outperformed both the BERT-based models and the pretrained Llama 3.1 baseline, achieving macro F1 scores of 0.992 on the validation set and 0.970 on the test set. PubMedBERT achieved the best performance among the BERT-based models, suggesting that pre-training on medical literature, making it better suited for understanding and processing medical texts, such as chest CT radiology reports. The effectiveness of [NUM] tokenization is likely attributed to its consistent numerical tokenization compared as compared to the standard BERT WordPiece tokenizer, which fragments numerical expressions requiring that all fragments be correctly tagged^33^. Among the Llama-based models, the instruction-tuned Llama 3.1 significantly outperformed the Llama 2 chat-tuned model and was marginally better than the Llama 3 instruction-tuned model. Meta attributes Llama 3.1’s superior performance to its enhanced reasoning capabilities and improved context length^29^. These improvements appear to have carried over in instruction-tuning, which allowed it to better handle the complexities of the dataset and achieve higher accuracy in extracting aortic measurements from chest CT radiology reports.

When applied to our extensive radiology report database, the model successfully extracted aortic measurements from 13.85% of reports, a rate consistent with both our labeled subset and prior work assessing aortic measurement reporting in CT radiology reports^40^. The extracted aortic measurements and dilation rates similarly aligned with findings from previous studies^41,42^. The resulting aortic measurement database is one of the largest of its kind, encompassing measurements from nearly 50,000 CT scans and over 28,000 patients, representing a valuable resource for advancing the study of aortic disease.

Initial enthusiasm for the potential of large language models (LLMs) in named entity recognition (NER) has recently been tempered. The expectation that general-domain LLMs could achieve domain-specific NER through in-context learning has been challenged by multiple studies, where fine-tuned BERT-based models consistently outperform LLMs^20,21,23^. Even with instruction-tuning, LLMs have, at best, matched the performance of BERT-based models—a disappointing outcome given their significantly larger parameter counts and the associated higher costs of fine-tuning and inference^23,25,26^. Researchers have suggested that the relatively poor NER performance of LLMs may stem from the limitations of their decoder-only transformer architecture and next-token prediction pretraining objective, compared to BERT’s encoder-only architecture and masked language modeling pretraining objective^43^. Our findings, however, challenge this hypothesis. In our study, the instruction-tuned Llama 3.1 achieved near-perfect performance on the NER task, surpassing the fine-tuned BERT-based models by what we consider a substantial margin. In addition to its fantastic performance, Llama 3.1 offers several additional advantages over BERT, including a much larger context length for analyzing longer text segments and more human-interpretable outputs, which streamline error analysis.

Despite these excellent results, additional work is needed to discern whether instruction-tuned generative LLMs have become the new gold standard for NER. Our findings are based on a single dataset, and the observed differences might be attributed to suboptimal fine-tuning of the BERT models, rather than the inherent superiority of the Llama 3 architecture. Further studies replicating these results across additional NER datasets is essential to substantiate these claims. Nonetheless, the ability of Llama 3 models to achieve this level of performance suggests that instruction-tuned generative LLMs hold significant promise for NER and could play a valuable role in clinical NER.

A significant advantage of our proposed methodology is its adaptability. The framework is agnostic to both the entities being extracted and the domain, enabling straightforward adaptations to various NER tasks. Instruction-tuning requires relatively few annotated samples, and open-source annotation tools such as Label Studio facilitate efficient, collaborative annotation processes. Frameworks like Hugging Face’s Transformers library offer well-developed pipelines for instruction-tuning general-domain LLMs, making them easily adaptable to diverse tasks. However, several barriers remain to the broader adoption of these techniques. LLMs still demand substantial computational resources for training and inference. For clinical projects, the additional requirement for HIPAA-compliant hardware introduces further costs and complexity. While existing pipelines are robust, they often require advanced coding and machine learning expertise, which may be beyond the scope of many clinical researchers. As the field of LLMs continues to evolve, these barriers are likely to diminish. Companies such as Microsoft and OpenAI are actively developing HIPAA-compliant implementations of their LLMs, and costs are expected to decrease as competition increases and the technology matures. If these trends persist, we anticipate that access to LLM instruction-tuning will become increasingly democratized, empowering clinical researchers to leverage these powerful tools.

Our study has several limitations. Both the validation and test sets are relatively small, with few annotations, making the results susceptible to variability as one or two errors can significantly impact model performance. Additionally, selecting a subset of sentences for inference may have led to the omission of relevant sentences when extracting measurements from the complete radiology report dataset. The BERT-based models used in our analysis are known to be sensitive to seed values^44^, which may have influenced their performance. Another limitation is the potential lack of generalizability to newly collected data. The validation and test sets share a temporal distribution with the training set, and medical data is prone to domain drift over time^45^. This could limit the applicability of our findings to datasets collected in different time periods or settings. We believe these limitations do not detract significantly from the overall value of our findings. Replication of our study in different datasets and settings is needed to validate our results and confirm the generalizability of our approach.

## Conclusion

In this study, we developed and evaluated a machine learning pipeline for extracting aortic measurements from chest CT radiology reports. The instruction-tuned Llama model achieved the best performance, surpassing state-of-the-art BERT-based models. Using this pipeline, we created a large, comprehensive database of aortic measurements from radiology reports, offering a valuable resource for aortic research. Our results highlight the potential of instruction-tuned generative LLMs in the NER domain, with a generalizable workflow that requires few labeled samples and modest computational resources. As the technology matures, this process is expected to become even more streamlined, enabling broader adoption in clinical research.

## Data Availability

All data produced in the present study are available upon reasonable request to the authors.

https://github.com/yalesurgeryresearch/RadTextExtractor/

## Acknowledgments

R.A. discloses support for the research of this work from the Yale Department of Surgery and the National Heart, Lung, and Blood Institute (NHLBI) of the National Institutes of Health (NIH) [grant number R01HL168473].

## Author Contributions

Conceptualization: EE, CSO, RA.

Data Curation: EE, SD, MT, AN.

Formal Analysis: EE.

Investigation: EE.

Methodology: EE, CSO.

Project Administration: EE, CSO.

Resources: EBS, CSO.

Supervision: RA, CSO.

Writing – Original Draft: EE, CSO.

Writing – Review & Editing: CSO, RA, PV, EBS.

All authors reviewed the manuscript.

## Data Availability

Study data are available upon reasonable request from the corresponding author, in accordance with institutional policies and any applicable data sharing or data use agreements.

## Code Availability

The code used in this study is open source and freely available under the MIT license. It can be accessed on GitHub at https://github.com/yalesurgeryresearch/RadTextExtractor/. This repository includes detailed documentation and examples to facilitate reproducibility and adaptation for related research.

